# CSF pharmacokinetics-pharmacodynamics of linezolid in critically brain injured patients, with or without central nervous system healthcare-associated infection. The PK-Pop-LCR Study: A Multicenter Pharmacokinetics and Pharmacodynamics Population Study

**DOI:** 10.1101/2024.12.13.24318990

**Authors:** Claire Dahyot-Fizelier, Alexia Chauzy, Kévin Chalard, Fanny Bernard, Hugues de Courson, Pierre-Etienne Leblanc, Gilles Francony, Russel Chabanne, Karim Lakhal, Raphaël Cinotti, Charles Gregoire, Julien Pottecher, Belaid Bouhemad, Assil Merlaud, Christophe Adier, Jean-Claude Lecron, Ombeline Remy, William Couet, Nicolas Gregoire, Sandrine Marchand, the PK-Pop-LCR Study Group

## Abstract

Linezolid is an alternative to vancomycin for treating Gram-positive central nervous system (CNS) healthcare-associated infections. The recommended dosing regimen remains debated. PK-Pop-LCR is a prospective population pharmacokinetic-pharmacodynamic multicenter study which included brain injured patients with an external ventricular drainage receiving linezolid at different dosing regimens. The cerebrospinal fluid (CSF) penetration of linezolid was investigated and a population pharmacokinetic model developed using plasma and CSF data. Monte Carlo simulations were conducted to calculate probability of target attainment (PTA) and cumulative fraction of response (CFR) in CSF against methicillin-resistant *Staphylococcus aureus* (MRSA) and methicillin-resistant *Staphylococcus epidermidis* (MRSE), for different dosing regimens. The plasma pharmacodynamic target, AUC/MIC > 100, was used in CSF.

Over 25 patients included, blind adjudication confirmed 14 cases of CNS infections. The mean AUC_CSF_/*f*AUC_plasma_ ratio was close to 80% with no difference between patients with and without CNS infection, despite higher CSF cytokines levels in CNS-infected patients. The recommended dose of 1200 mg/24h allowed to reach PTAs ≥ 90% only for MICs ≤ 0.5 mg/L, and CFR of 3.2% and 40% for MRSA and MRSE, respectively. 2 700 mg/24h would allow to achieve PTA > 90% for MIC up to 1 mg/L and CFR of 90% for MRSE but none of dosing regimens tested was appropriate for MRSA infections.

We confirmed the extensive CSF distribution of linezolid. Higher doses than those recommended should be considered to treat CNS infection in critically brain injured patients. However, pharmacodynamic target for CNS infections should be further investigated to confirm these findings.

**Fundings:** French Ministry of Health.

## INTRODUCTION

Central nervous system (CNS) healthcare-associated infections (CNS-HAI) are associated with high morbidity and mortality. Recently, a mortality rate of 9% was reported, with an unfavorable prognosis in 50% of surviving patients. One year mortality rises to almost 40% in ventriculitis^1,2^. In literature, incidences varies according to the clinical situation, ranging from less than 1% in post-operative cases of scheduled surgery to 10% in average in external ventricular drain (EVD) associated infections in ICU^3^.

In two-thirds of cases, causative organisms are Gram-positive bacteria, with a majority of *Staphylococcus aureus* and coagulase-negative bacteria^4^. To date, high-dose of vancomycin is still recommended as a first-line for CNS-HAI, despite its limited CNS distribution and narrow therapeutic index due to its renal toxicity^5^. Indeed, physiological barriers in the brain, blood-brain barrier (BBB) and blood-cerebrospinal fluid barrier (BCSFB) limit penetration of hydrophilic antibiotics such as vancomycin, exposing patients to the risk of therapeutic failure. Recently, vancomycin diffusion was reported to be between 9 and 13% in patients with CNS infections, with median CSF concentrations of 2.9 +/- 2.2 mg/L ^6^, failing to reach the EUCAST minimum inhibitory concentration (MIC) threshold for resistant *Staphylococcus*^7^.

Linezolid, an oxazolidinone agent, is indicated against multidrug resistant Gram-positive bacteria for the treatment of community- and hospital-acquired pneumonia and skin structure infections^8^. It is also increasingly prescribed for CNS infections in ICU patients as an alternative when methicillin-resistant *S. aureus* (MRSA) strains have a vancomycin minimum inhibitory concentration (MIC) higher than 1 mg/L, as linezolid characteristics enable it crossing barriers more easily with less systemic adverse events^5^. More recently, linezolid was proposed in tuberculous meningitis, especially for drug-resistant ones as data on CNS penetration of new and repurposed anti-tuberculosis drugs, are very limited ^9–11^. Indeed, high linezolid CSF penetration from 60 to 90% was reported, but mainly in non-infected situations or extra-cerebral infections^12^. However, those few studies have limitations: few patients, no proven CNS infection for most of them, comparison of CSF linezolid concentrations to total plasma concentrations, no inflammatory biomarkers and EVD flow rates reported, and only one administration scheme studied ^9,13–18^. Despite high CNS concentrations of linezolid, several studies reported that pharmacodynamics (PD) targets were not achieved, suggesting the need to increase the studied recommended doses of 600mg q12h^13,14^.

The aim of the present study was to explore the linezolid CSF pharmacokinetics (PK) and theoretical efficacy in ICU patients with an EVD, at different dosing regimens, with or without infection.

## METHODS

### Study design and participants

The PKpopLCR study is a national prospective population PK/PD multicenter study (*CPP17-016a/2017-002993-37*) of 9 broad-spectrum antibiotics involving a total of 14 French ICU. It was approved by the Committee for the Protection of Persons (CPP) of Sud-Ouest and Outre-Mer IV (CPP Sud-Ouest et Outre-Mer IV; EudraCT: 2017-002993-37) and received the authorization of “Agence Nationale de Sécurité du Médicament et des Produits de Santé” (ANSM). In this article, we reported the PK/PD results of linezolid, a substudy nested within the PKpopLCR study, conducted in eleven of the participating centers.

Brain injured patients (≥18 years of age) of both sexes and all ethnic groups, requiring intensive care management with EVD, and treated for a CNS or extra-CNS infection by linezolid were eligible to the trial. All declared cases of CNS infection were reviewed by a central and masked adjudication committee, composed of two senior intensivists, who had access to all anonymized monitored data to confirm the CNS infections according to IDSA definition^5^. Patients were recruited from January 2019 to February 2020. Written informed consent was obtained from legal surrogate before inclusion as patients were in coma.

Exclusion criteria were acute renal failure defined with a creatinine clearance (Clcr) < 50 mL/min and/or under continuous hemodialysis, pregnancy, contraindication to linezolid, patient or family refusal to be involved in the study and patient with reinforced protection or deprived of freedom subsequent to a legal or administrative decision. Patient clinical data were recorded on a data collection template by reviewing electronic medical records, including identification of the pathogen and MIC, linezolid dosing regimen, number of injections before the PK study; demographics, such as age, gender and actual body weight (WT); routine blood and biochemical indexes at the inclusion. On each day of PK sampling, following data were collected in plasma: creatinine clearance, glucose, albumin, protein and leukocytes and, in CSF: proteins, lactates, glucose, albumin and bacteriological analysis; CSF volume collected over the 24 hours was reported and SOFA score was calculated.

### Intervention and sample collection

All patients received linezolid (infusion solution at 2mg/mL) by intravenous (IV) infusion over 30 minutes, 600 mg twice a day (b.i.d., n=17 patients) or 600 mg thrice a day (t.i.d., n=8). The PK study was performed within a dosing interval at steady state.

Blood samples were collected at the start of IV injection (H0), at the end of infusion (H0.5), 2 hours and 6 hours after the administration, and just before the next administration, i.e. 8 or 12 hours after administration depending on the dosing regimen (t.i.d. or b.i.d. respectively). Each CSF sample was collected over a one–hour interval *via* the EVD under aseptic conditions, starting the hour before the beginning of the infusion (H-1-H0), then from the beginning of the infusion to the first hour (H0-H1) and between H1-H2, H2-H3, H3-H4, H5-H6 whatever the dose administered. A last CSF sample was collected from H7 to H8 or from H11 to H12 depending on the dosing regimen, t.i.d. or b.i.d. respectively. On the day of PK, one additional blood and CSF sample was collected for albumin, Interleukin (IL) 6, IL8, IL10, IL17 and TNF-alpha measurements. Blood samples were centrifuged at 3000rpm (4°C; 10min). Plasma and CSF samples were stored at -80°C until analysis. The exact time of collection of each sample was recorded and the dead volume of the EVD tubing was taken into account for PK analysis.

### Linezolid Assay

Total plasma and unbound CSF concentrations of linezolid were measured using an appropriate validated LC– MS/MS method with a limit of quantification (LOQ) of 0.01 mg/L for both matrices (Supplemental Materials).

### Cytokine and albumin measurements

The concentrations of IL6, IL8, IL10, IL17A and TNF-alpha were quantified in plasma and CSF with the Luminex 200™ plateform (Luminex Xmap Technology) coupled with xPONENT™ software by using the MILLIPLEX MAP Human Cytokine/Chemokine magnetic bead panel kit (Millipore Corporation, Billerica, MA) according to the manufacturer’s instructions. Some samples were assayed in duplicates to validate assays. Albumin concentrations in plasma and CSF were quantified by immunonephelemetry assay using an Atellica^®^ analyzer (Siemens). CSF-to-plasma albumin concentrations ratios (Q_alb_) were then calculated to assess the BCSFB function. Values between 7 and 10 × 10^-3^ indicated a mild dysfunction, those between 10 and 20 × 10^-3^ a moderate dysfunction and those above 20 × 10^-3^ a severe dysfunction of BCSFB^19^.

### PK/PD analysis

#### Population PK analysis

Total plasma and CSF concentrations of linezolid were analyzed simultaneously using the non-linear mixed-effect modelling approach in NONMEM 7.4 (ICON Development Solutions, Ellicott City, MD, USA). All model estimations were conducted using the first order conditional estimation method with interaction (FOCE INTER). A detailed description of the development, evaluation and the covariate selection is available as Supplementary data. Briefly, the structural model consisted of one compartment with linear elimination for plasma, one compartment for ventricular CSF and one compartment for CSF obtained by EVD (Figure S1). As only the unbound fraction of the antibiotic can distribute into and out of the CSF, predicted total plasma concentrations were converted to free concentrations, assuming a protein binding of 31%, before fitting CSF data^8^, while protein binding of linezolid in CSF was considered negligible due to the low concentrations of blood-derived proteins, particularly albumin, in CSF^20^. The bidirectional passage across the BCSFB was characterized by a clearance into the CSF (Q_CSF,in_) and a clearance out of the CSF (Q_CSF,out_). Elimination of linezolid from the CSF via the EVD was taken into account by fixing the flow rate of the EVD (CL_EVD_) for each patient to its actual value. The volume of CSF collected at each time interval by the EVD (V_EVD_) was determined experimentally and used as a fixed value in the model. The influence of various covariates such as creatinine clearance, weight and inflammatory biomarkers on the PK parameters of the structural model was evaluated (Supplemental materials). The free maximal concentrations (C_max_) and the free areas under the concentration-time curves from 0 to 24h (*f*AUC_0-24_) in plasma and CSF were estimated at steady-state from 5 000 concentration-time profiles simulated with the final model.

#### Probability of Target Attainment (PTA) and Cumulative Fraction of Response (CFR)

The probability of achieving the PK/PD efficacy target (*f*AUC_0-24_/MIC > 100) of linezolid in plasma and CSF was estimated from Monte-Carlo simulations for 1 200, 1 800 and 2 700 mg daily doses, delivered in 2 or 3 injections (over 30 or 180 min) or by continuous infusion. The PD parameter *f*AUC_0-24_/MIC > 100 was chosen according to the single clinical study which reported the optimal PD parameter to predict infections’ cure in seriously infected patients^21^, and as it is the most often retained in recent literature ^13,16,17^. No study explored the optimal PD parameter in CNS infections. Linezolid concentration–time profiles of 5 000 patients were simulated for each dosing regimen with the final model. To evaluate whether the proposed linezolid dosing regimens achieved adequate exposures for maintaining efficacy against methicillin resistant *S. aureus* (MRSA) and *S. epidermidis* (MRSE) isolates encountered in clinical practice, PTAs were compared with corresponding linezolid MIC frequency distributions from EUCAST^22^. CFRs were calculated for each dosing regimen^23^.

### Statistical analysis

All statistical analysis was performed using R statistical software (v4.2.3; R Core Team 2023). Descriptive statistics were presented as median and interquartiles. Cytokines concentrations between plasma and CSF, Q_alb_ and cytokine concentrations in CSF between CNS and extra-CNS infection groups were compared with a nonparametric Mann Whitney U test (p < 0.05). Cytokines concentrations below the detection limit were considered as half of this threshold.

## RESULTS

### Patients and data

Among the 25 patients included, 16 CNS (11 associated-healthcare meningitis and 5 ventriculitis) and 9 extra-CNS (8 Ventilator-Associated Pneumonia and 1 pulmonary abscess) infections were declared. Among the 16 CNS infections, 14 were confirmed after adjudication. The most frequent pathogens identified at the infection site were *Staphylococcus epidermidis* (7/14), then in decreasing order *Cutibacterium acnes* (3/14), *Staphylococcus aureus* (1/14), *Staphylococcus pettenkoferi* (1/14), *mycobacterium tuberculosis* (1/14) and 1 ventriculitis with a various flora of Gram-negative bacillus. Patients’ demographics, clinical, biological characteristics and pathogens identified are summarized in Table S1. The underlying diseases were mostly hemorrhagic stroke, with subarachnoid or intracranial hemorrhage. A total of 87 plasma and 159 CSF samples were considered for PK analysis.

### Inflammatory markers

In the “CNS Infection” group, median concentrations of all cytokines except TNF-alpha were significantly higher in CSF than in plasma (Figure 1). No difference was observed in the “Extra-CNS Infection” group.

**Figure 1:**
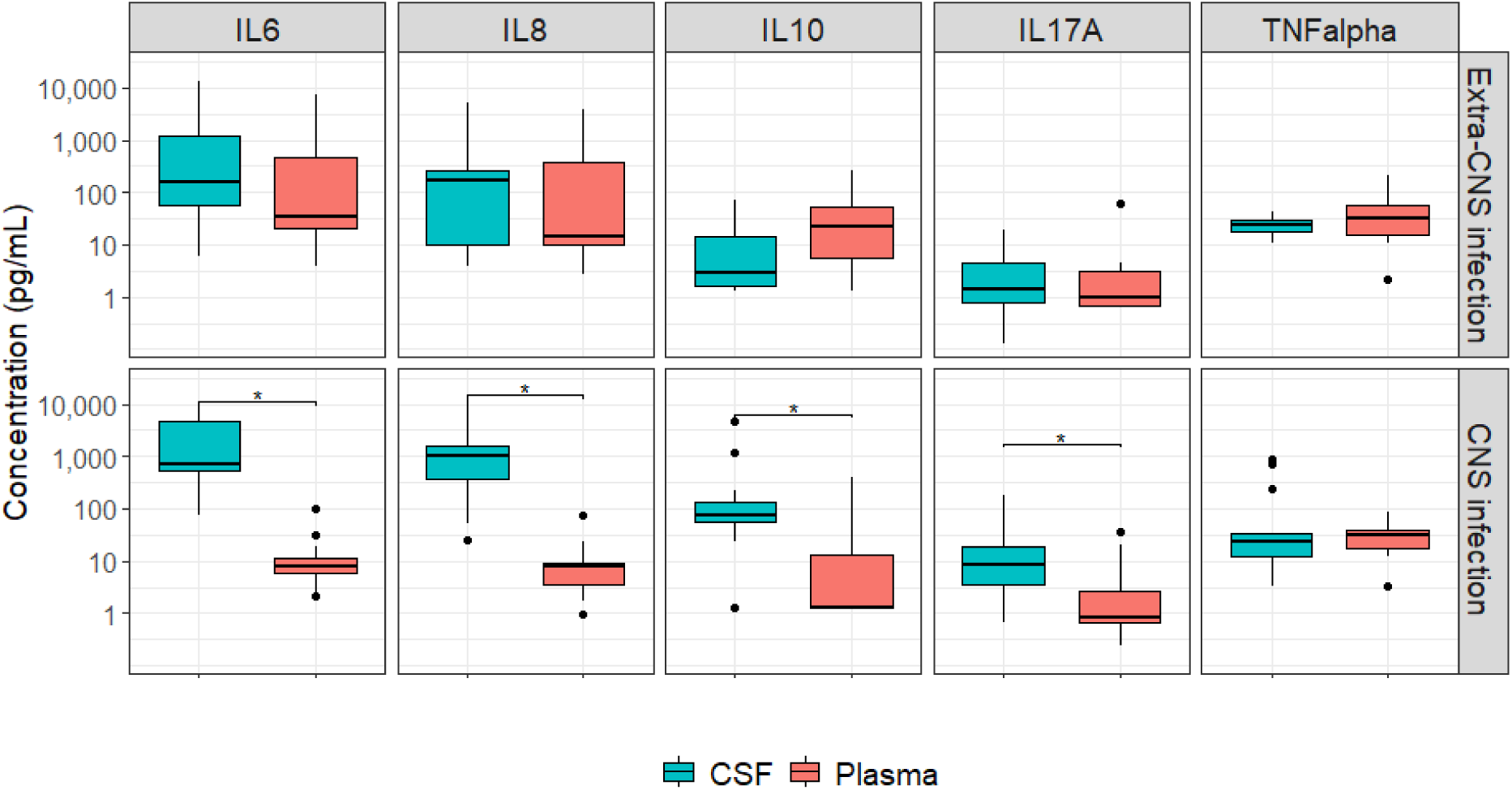
Boxplots of concentrations of IL6, IL8, IL10, IL17A and TNF-alpha in CSF (in blue) and plasma (in red) for “Extra-CNS Infection” group (Top line boxplots) and for “CNS Infection” group (Bottom line boxplots). * p<0.05.

When comparing the median concentrations of IL in the CSF between the two groups of patients, a significant difference was observed only for IL10 and IL17A (Figure 2). However, although not statistically significant, a 4-fold increase of median IL6 concentration and 6-fold increase of median IL8 concentration were observed in patients with CNS infections compared to patients with extra-CNS infections (726 pg/mL [522-4823] vs 161 pg/mL [59-1195] and 1016 pg/mL [441-1553] vs 172 pg/mL [10-266], respectively).

**Figure 2:**
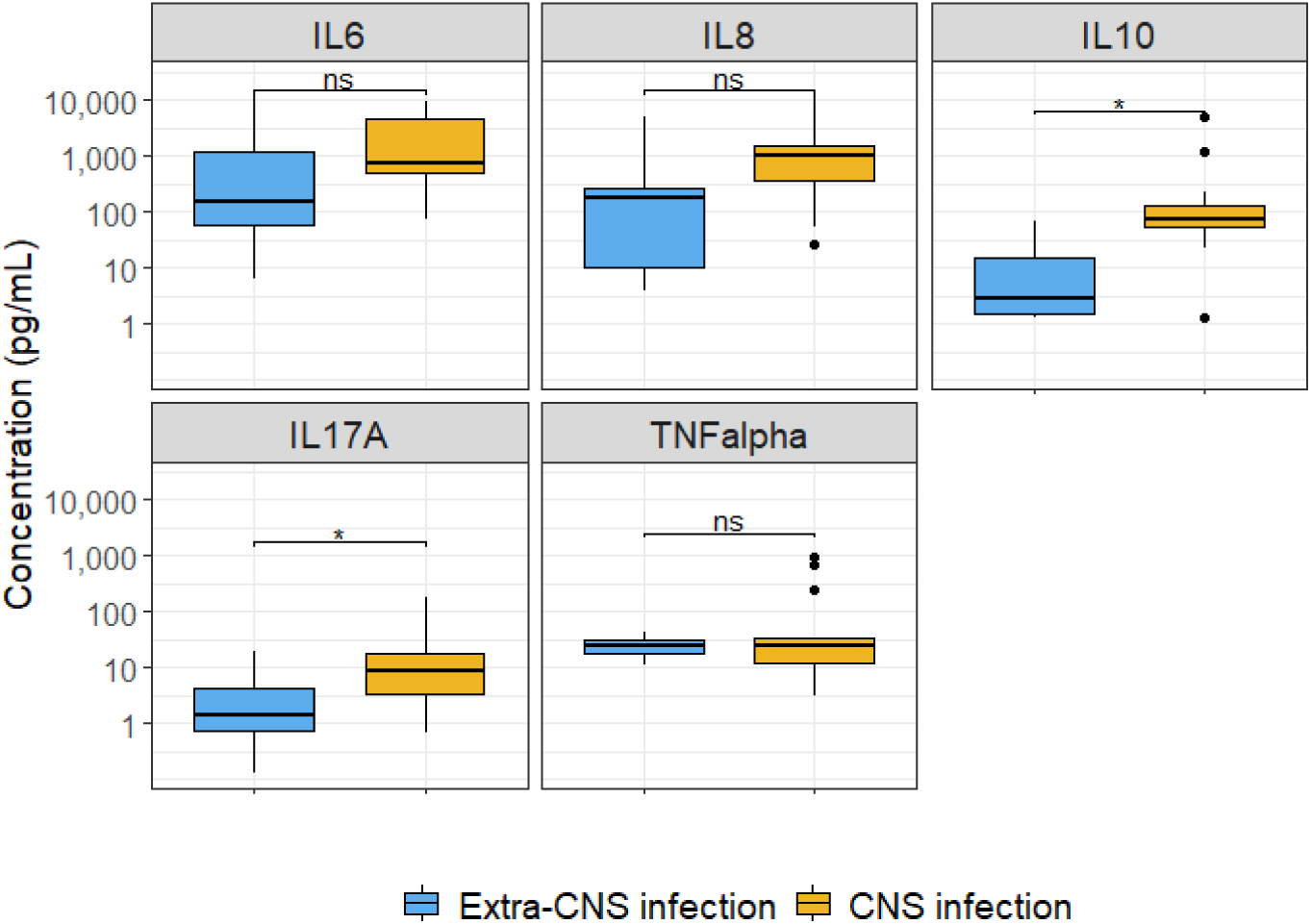
Boxplots of CSF concentrations of IL6, IL8, IL10, IL17A and TNF-alpha in patients with extra-CNS infections (in blue) and patients with CNS infections (in yellow). * p<0.05.

The median Q_alb_ was not significantly different between “CNS Infection” (7.3 × 10^-3^ [5.5 × 10^-3^ - 18.4 × 10^-3^]) and “Extra-CNS Infection” (8.9 × 10^-3^ [6.3 × 10^-3^ - 14.2 × 10^-3^]) groups, with a larger variability in the “CNS Infection” group (Figure S2). Two patients exhibited severe dysfunction, both belonging to the “CNS Infection” group, while all others showed moderate or mild dysfunction.

### Plasma and CSF concentrations and population PK model parameters

The final PK model described the observed data in plasma and CSF well, as indicated by the visual predictive check (VPCs), individual predictions vs. observed data and goodness of fit plots (Figures S3, S4, and S5). The final model parameters were all estimated with good precision (Table S2).

Predicted free plasma and CSF concentrations for the two dosing regimens are shown on Figure 3. Compared with plasma, the predicted C_max_ (mean ± SD) in CSF was about two times lower whatever the dose (Table 1), but also smoother and delayed (Figure 3, Table 1). However, the predicted *f*AUC_0-24_ in plasma and CSF were similar with mean linezolid CSF-to-plasma *f*AUC_0-24_ ratios close to 80% for both dosing regimens (Table 1).

**Figure 3.**
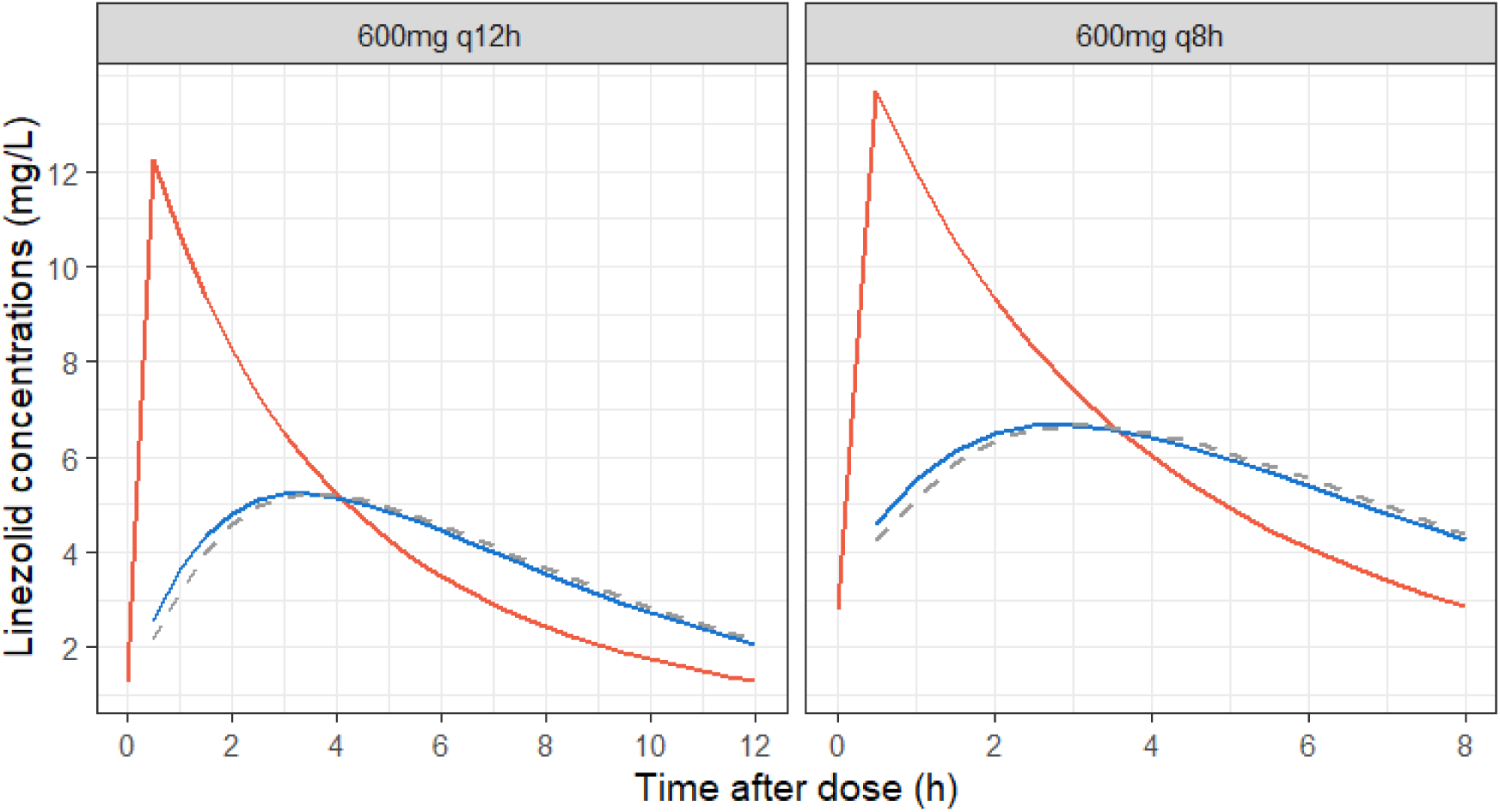
Simulations of the mean free concentration-time profiles of linezolid at steady-state in plasma (red solid lines), ventricular CSF (blue solid lines) and EVD (grey dotted lines) after administration of 600mg q12h and 600mg q8h.

**Table 1.** Median free plasma and CSF PK parameters at steady-state after administration of 600mg q12h (n=17 patients) and 600mg q8h (n=8 patients) of linezolid.

|  | 600 mg q12h |  | 600 mg q8h |  |
| --- | --- | --- | --- | --- |
|  | Plasma | CSF | Plasma | CSF |
| **Free $C_{\max}$ (mg/L)** | 12.2 ± 3.4 | 5.3 ± 2.0 | 13.7 ± 3.7 | 6.7 ± 2.7 |
| **$T_{\max}$ (h)** | 0.5 ± 0.0 | 3.4 ± 0.6 | 0.5 ± 0.0 | 2.9 ± 0.4 |
| **Free $AUC_{0-24}$ (h.mg/L)** | 102.4 ± 42.6 | 85.3 ± 37.6 | 150.7 ± 61.3 | 123.3 ± 53.6 |
| **$AUC_{CSF}/fAUC_{pl}$** | 0.838 ± 0.151 | | 0.823 ± 0.147 | |

Over all relevant covariates, only the effect of body weight on the central plasma compartment volume of distribution was found to significantly improve the model. Thus, neither CNS infection nor inflammation biomarkers and Q_alb_ had a significant impact on linezolid CSF distribution and were therefore not included in the final model. However, linezolid elimination rate from CSF via the EVD (CL_EVD_) was not negligible, ranging between 2.51 and 20.17 mL/h (Table S1), corresponding to 27.8% of Q_CSF,out_ on average, which tended to decrease the mean CSF linezolid exposure, resulting in a lower CSF-to-plasma *f*AUC_0-24_ ratio as CL_EVD_ increased (Figure 4).

**Figure 4:**
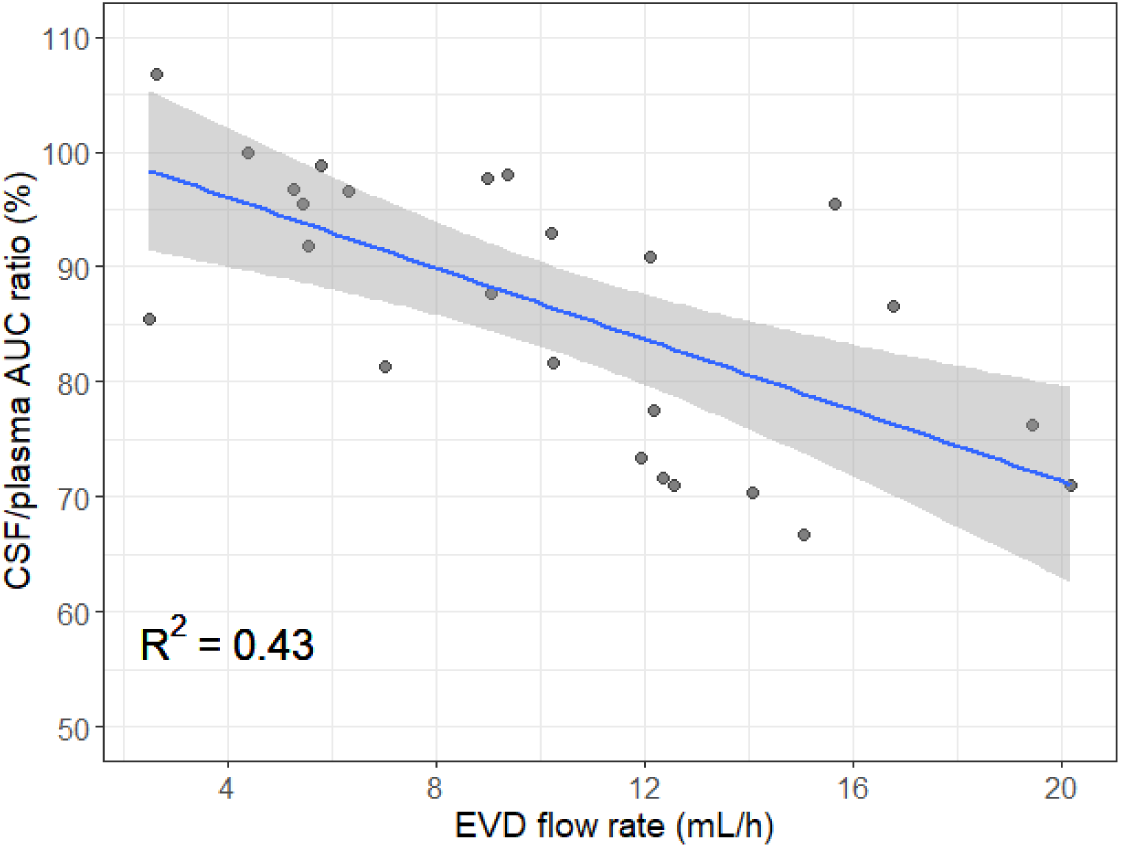
Relationship between CSF/free plasma AUC ratio and EVD flow rate. Circles represent the individual predictions of CSF/free plasma AUC ratio for the mean EVD flow rate measured on the PK day for each patient. The line represents the fit of a linear function and the grey shaded area the 95% confidence interval.

### PTA and CFR

The probability of reaching *f*AUC_0-24_ ⁄ MIC values higher than 100 for various dosing regimens are shown on Figure 5, and the CFRs for the treatment of MRSA and MRSE in Table 2. The predicted PTAs and consequently the CFRs were similar between plasma and CSF. The usually recommended daily dose (1 200 mg/24h) would allow reaching PTA ≥ 90%, only for MIC ≤ 0.5 mg/L and CFR of 3.2% and 40% for MRSA and MRSE, respectively. Increasing the daily dose to 1 800 mg would result in higher PTA values, but still insufficient, as the PK/PD target would be only reached in 72.5 % of simulated patients for an MIC = 1 mg/L. Only a daily dose of 2 700 mg would allow to achieve PTA > 90% against pathogens with MIC up to 1 mg/L and thus cover the MICs frequency distribution for MRSE with CFR = 90%. None of the dosing regimens tested appear to be appropriate to treat MRSA infections (CFR ≤ 32%). None of the tested modalities of administration of linezolid, thrice daily rather than twice, or continuous infusion instead of intermittent, had any impact on PTA and CFR values.

**Figure 5.**
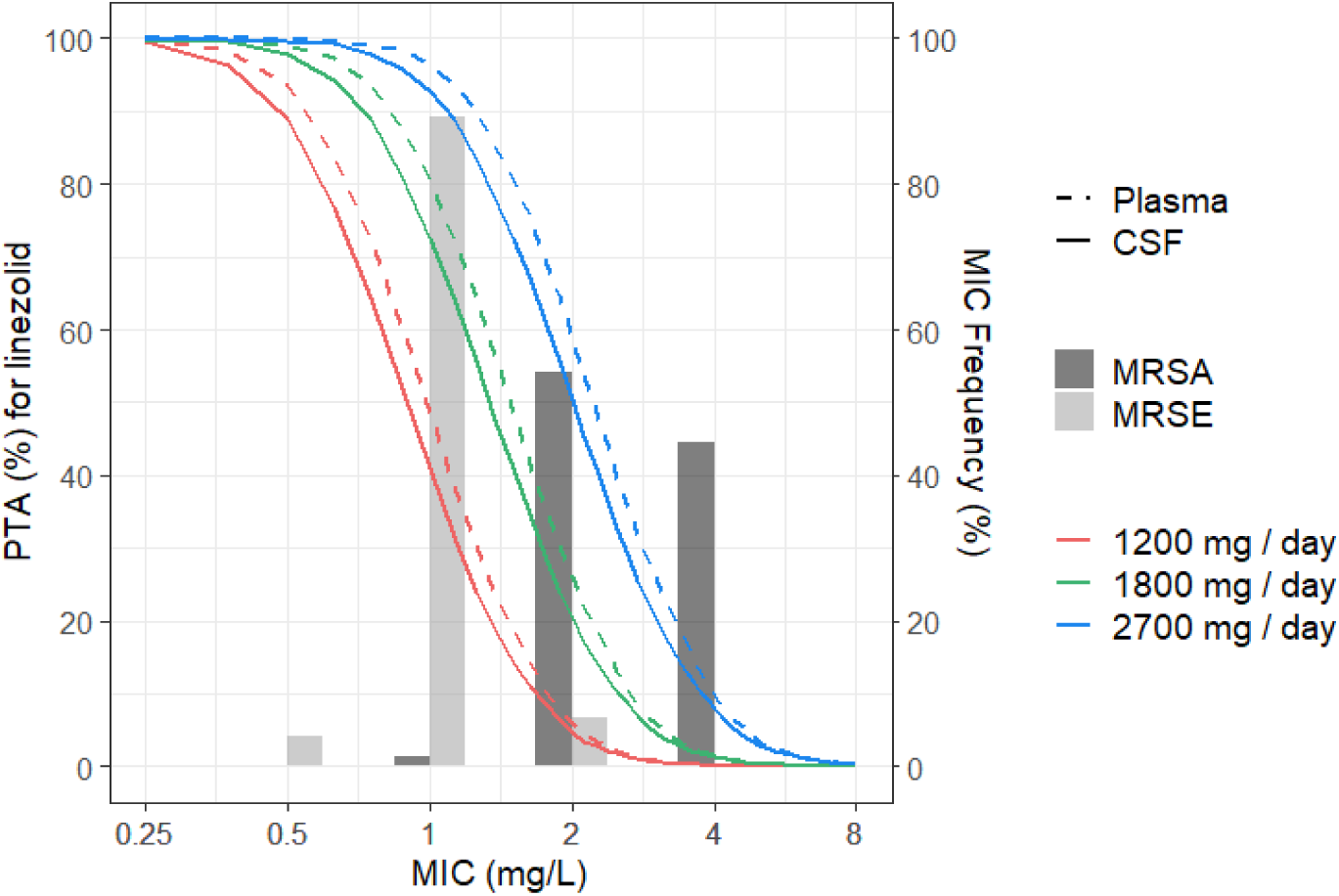
PTA (*f*AUC_0-24_ > 100) in plasma (dashed lines) and CSF (solid lines) in simulated patients receiving different daily doses of linezolid (1 200, 1 800 and 2 700 mg/day). The distribution of linezolid MICs for methicillin resistant *S. aureus* (MRSA) and *S. epidermidis* (MRSE) is shown.

**Table 2.** Cumulative fraction of response (CFR) of linezolid against methicillin resistant *S. aureus* (MRSA) and *S. epidermidis* (MRSE) in simulated patients receiving different daily doses.

|  |  | 1 200 mg/day | 1 800 mg/day | 2 700 mg/day |
| --- | --- | --- | --- | --- |
| MRSA | Plasma | 3.9% | 16% | 38% |
|  | CSF | 3.2% | 13% | 32% |
| MRSE | Plasma | 48% | 78% | 94% |
|  | CSF | 40% | 70% | 90% |

## DISCUSSION

To our knowledge, this is the most comprehensive population PK study of linezolid in plasma and CSF in a large number of critically ill patients with proven CNS infection, which explored several linezolid dosing regimens.

Few studies conducted a robust PK analysis to describe linezolid disposition in CNS, with heterogeneous methodological approaches and small sample sizes, preventing accurate characterization of the wide interindividual variability in the ICU population^24^. Traditional compartmental modelling was only occasionally conducted to characterize the distribution of linezolid in CSF ^13^. In the present study, given the low accumulation upon linezolid repeated doses, the estimated mean steady-state total volume of distribution (Vd) (36.4 L) was in line with those previously reported in similar patients groups, at steady state (42.8 L, 40.1 L and 52.8 L) and also after the first administration (48.7 L and 77.3 L)^14–18^. The same observation can be made regarding clearance, the total clearance of linezolid (8.4.L/h) in the present population being similar to that reported in the previously mentioned studies^14–18^. Only one study reported higher findings without any clear explanation (Vd=101.3 L; CL=16.6 L/h)^13^.

Considering linezolid CSF distribution, flat concentrations profiles were observed with around 3 hours delay in peak concentrations compared to plasma, in accordance with the amphiphilic properties of linezolid and with previous reported observations in ICU patients^13–17^. Our results confirmed a good central penetration of linezolid around 80%, despite significant inter-studies variability with CSF/plasma AUC ratio ranging from 41.3% to 77% in patients without CNS infection^13,15–18^, and from 80% to 98% in patients with CNS infections^14,25^. However, all these previous studies compared AUCs in CSF with total plasma AUCs, without taking into account the protein binding of linezolid, probably leading to a slight underestimation of ratios.

In our study, we observed significantly higher CSF than plasma concentrations of all cytokines, except for TNF-alpha, in patients with CNS infection which suggest a local neuroinflammation in these patients. To our knowledge, this difference of response in serum and CSF between ICU patients with or without CNS infection was not previously reported. In bacterial meningitis, an increase of all cytokines in CNS was already described, but only transiently for TNF-alpha and IL-1^26^. However, despite these significant differences, cytokine levels were not identified as covariates influencing linezolid diffusion, unlike meropenem for which high CSF IL-6 levels were associated with increased CSF penetration in ventriculitis patients^27^. Besides, the brain barriers integrity marker, CSF/plasma albumin ratio (Q_alb_), did not influence linezolid diffusion either, contrasting with the impact described for meropenem and vancomycin, known for their limited CNS diffusion^28,29^. In our population, median Q_alb_ below 10 × 10^-3^ in both groups (Figure S1) suggested only a mild BCSFB dysfunction^19^, which could explain the lack of influence of this covariate on linezolid diffusion, whose penetration is particularly good. Nevertheless, caution must be taken when interpreting this parameter and its influence in CNS infections studies as it may be influenced by several factors, including the rostrocaudal gradient within the CSF, with Q_alb_ twice as low in cerebral ventricles samples as in lumbar ones^30,31^. Interestingly, the EVD rate influenced the CSF exposure of linezolid, as also recently described for vancomycin and meropenem in literature^32,33^. The higher the EVD rate, the lower the AUC ratio (Figure 4), reflecting the increased linezolid clearance. Thus, in case of CNS infection, it might be advisable to limit CSF drainage, under optimal cerebral hemodynamics, to limit the additional clearance due to EVD, reducing CSF linezolid concentrations.

From a pharmacodynamics perspective, plasma PK/PD index related to linezolid efficacy is not clearly defined and varies among studies, utilizing either AUC/MIC or %T>MIC^13–18^. In the first study describing *in vivo* linezolid PD from animal models of acute infections, the more relevant of the two PK/PD parameters was difficult to determine^34^. In seriously ill patients, a linear relationship between these two parameters was characterized for linezolid at AUC/MIC values < 120, and higher clinical success rates occurred at AUC/MIC ratios of 80-120 or T>MIC of 100%^21^. However, careful interpretation of these results is essential as only 50% of patients received linezolid intravenously and only 2 patients were treated for a CNS infection. In literature, plasma PK/PD targets are currently used to optimize the treatment of cerebral infections, with AUC/MIC > 100 most often retained, as no target based on CSF concentrations exist^13,16,17^. Thus, depending on studies, indexes and targets used varied, leading to different conclusions. Luque *et al.,* targeting 24-h AUC/MIC ≥ 80 in CSF, considered that higher than standard doses were necessary (AUC/MIC of 21 for pathogens with a MIC of 2 mg/L)^13^. In contrast, Beer *et al.* suggested that 600 mg linezolid twice daily provided satisfactory PK/PD results in ventriculitis patients (T>MIC of 99.8% for pathogens with a MIC of 2 mg/L)^14^.

According to our results, selecting as PK/PD target a 24-h CSF AUC/MIC ratio > 100, the recommended dose of 1200 mg/day achieved PTAs > 90% only for staphylococci with MICs < 0.5 mg/L. Increasing daily dose to 2700 mg/day would allow reaching PTA > 90% for MICs up to 1 mg/L only, and would allow covering linezolid MIC distribution for MRSE (CFR = 90%) but not for MRSA (CFR = 32%). Whatever doses, none of the administration modalities tested showed any impact on PTA and CFR values, and hence any benefit in terms of efficacy for MRSA-infected patients. These results could challenge linezolid indication, in case of a bacteria with a MIC greater than 1 mg/L considering that linezolid is recommended as a second-line treatment or for MRSA with vancomycin MIC higher than 1 mg/L^5^. Nevertheless, its clinical efficacy has been reported in literature in cases of failure of vancomycin treatment for MRSA infections, including infections of shunts, with the recommended 600mg twice daily, whereas most PK/PD studies in CSF recommend increasing dosages^35^.

If we had adopted a CSF PK/PD target of 100% *f*T>MIC, results would have diverged significantly. While a daily dose of 1800 mg seemed inadequate for treating pathogens with MICs exceeding 0.5 mg/L when targeting CSF AUC/MI > 100 (PTAs = 98%, 73% and 21% for MICs of 0.5, 1 and 2 mg/L respectively), and regardless of dose fractionation (Figure 6), it seems that if the chosen target had been 100% CSF *f*T>MIC, then splitting the dose to 600 mg three times a day or opting for continuous infusion (1800 mg/day) would have resulted in higher PTA values for pathogens with an MIC up to 2 mg/L (≥ 85% and ≥ 98%, respectively)(Figure S6). Thus, doses lower than 2700 mg/day would have been proposed to treat MRSE and MRSA infections. For MRSE, a daily dose of 1200 mg by continuous infusion would have been sufficient to achieve CFR > 95%, while a daily dose of 2400 mg administered as a continuous infusion would have been theoretically more appropriate for MRSA (CFR = 95%) (Table S3).

Increasing dosing regimens raises concerns about the risk of linezolid hematological toxicity. However, an increased risk was exclusively reported in cases of renal failure, with the need to adapt dosing regimen when Clcr < 60 mL/min^36,37^. In our population, the majority of patients had an augmented renal clearance (ARC) (18/25, Table S1) and none of them exhibited any residual concentration over 10 mg/L, i.e. the threshold of toxicity, even when they received 1800 mg per day ^38^. Recent studies involving ICU patients highlighted risks of under-dosing in cases of ARC, indicating the usefulness of dose adaptation. An ICU prospective multicenter study involving 152 intensive care patients recommended increasing linezolid up to 2400 daily for patients with ARC^39^. An early therapeutic drug monitoring is perhaps the first step to optimize linezolid dosages in ICU patients, even if linezolid does not require adjustment based on renal clearance theoretically.

The first strength of our study is its design. Its multicenter framework allowed to include the highest number of patients with cerebral infections, all confirmed by a blinded adjudication committee (n=14), and to evaluate linezolid cerebral penetration at two dosing regimens (600mg b.i.d for 17 patients; 600mg t.i.d for 8 patients). Second, it is the largest ICU PK/PD study based on Monte Carlo simulations reporting PTA and CFR of linezolid against MRSE and MRSA in critically ill patients. Additionally, cytokine concentrations in the CSF and plasma in both patient groups were measured. Although no impact of inflammation on linezolid distribution into CSF was observed between CNS infected patients and extra-CNS infected patients, this difference in cytokine behavior based on infection type in intensive care patients was not shown previously. For the first time also, the flow rate of the EVD was described to influence linezolid concentrations in CSF.

The first limitation of our study is the lack of patients with renal clearance lower than 80 mL/min, making it impossible to demonstrate any link between linezolid and creatinine clearances, and to propose a dosing regimen adjustment based on renal function. Secondly, the absence of follow-up of clinical recovery and bacterial eradication made impossible to determine a PD threshold adapted to CNS infections. The need for targets based on CSF concentrations is urgent, but the complexity of gathering a large number of infected patients undergoing monotherapy treatment as clinicians often resort to antibiotic combinations, make PK/PD analysis difficult and uncertain^40^. An approach based on a hollow-fiber infection model that enables continuous culture of bacteria and mimics the clinical drugs concentration profiles in CSF, could overcome these limitations.

In conclusion, study findings support extensive linezolid diffusion into CSF of severely brain-injured patients, influenced by the EVD flow rate, and highlight that currently recommended doses (600mg b.i.d) would not be appropriate for covering infections due to staphylococci in ICU patients with normal or ARC, whatever PK/PD index chosen. Considering AUC/MIC > 100 in CSF, 2700mg daily would cover only MRSE, whereas with 100% *f*T>MIC, daily continuous infusions of 1200mg and 2400mg would be appropriate for MRSE and MRSA, respectively. However, these results should be interpreted with caution without any well-defined PD target for cerebro-meningeal infections. Further PK/PD studies including clinical outcomes in larger population are necessary to identify such linezolid PD index in CSF of severe brain-injured patients.

## Data Availability

All data produced in the present study are available upon reasonable request to the authors.

## Acknowledgments

We thank the patients and their relatives and caregivers, local site clinical and research staff, regional coordinating center staff of the university hospital of Poitiers, and all the members of the PK-Pop-LCR Study for their active participation.

## Authors contributions

CDF, AC, NG, WC and SM conceived the study, participated in protocol development, study design, data interpretation, and co-wrote the first draft of the report. CD-F coordinated the study. CDF, FB, KC, HdC, PEL, GF, RCh, RCi, JP, BB, AM participated in inclusion of patients and data collection. DCF and FB participated in the masked central adjudication committee. OR verified and CA, JCL analyzed samples. CDF, OR, AC, JCL, FB and AM accessed and verified the data. AC, AM, NG and SM provided statistical and pharmacokinetic expertise. All authors attest to have had full access to all the data in the study, contributed to manuscript revision and to have approved the final manuscript prior to submission. All collaborators for the PK-Pop-LCR study study group, full names and academic degrees, are reported in the supplementary appendix.

## Further Members of the PK-Pop-LCR study group with linked authorship in Pubmed

Fanny BERNARD, M.D (Centre Hospitalier Universitaire de Poitiers, Service d’Anesthésie-Réanimation et Médecine Péri-Opératoire, Poitiers, France.), Carine DOMINIQUE, M.D (Centre Hospitalier Universitaire de Poitiers, Service d’Anesthésie-Réanimation et Médecine Péri-Opératoire, Poitiers, France.), Tony TRIQUARD, M.D (Centre Hospitalier Universitaire de Poitiers, Service d’Anesthésie-Réanimation et Médecine PériOpératoire, Poitiers, France.), Amélie RUBIN, M.D (Centre Hospitalier Universitaire de Poitiers, Service d’Anesthésie-Réanimation et Médecine Péri-Opératoire, Poitiers, France.), Aude PERALEZ, M.D (Centre Hospitalier Universitaire de Poitiers, Service d’Anesthésie-Réanimation et Médecine Péri-Opératoire, Poitiers, France.), Jean-Claude LECRON, M.D., PhD (Université de Poitiers, LITEC, France), Adriana DELWAIL (Université de Poitiers, LITEC, France), Rémy BELLIER, M.D. (Centre Hospitalier Universitaire de Poitiers, Service d’Anesthésie-Réanimation et Médecine Péri-Opératoire, Poitiers, France.), Thierry BENARD, M.D. (Centre Hospitalier Universitaire de Poitiers, Service d’Anesthésie-Réanimation et Médecine Péri-Opératoire, Poitiers, France.), Nadia IMZI, C.R.A. (Centre Hospitalier Universitaire de Poitiers, Direction de la Recherche Clinique, Poitiers, France.), Angela KOSTENCOVSKA, C.R.A (Centre Hospitalier Universitaire de Poitiers, Direction de la Recherche Clinique, Poitiers, France.), Sabrina SEGUIN, C.R.A (Centre Hospitalier Universitaire de Poitiers, Direction de la Recherche Clinique, Poitiers, France.), Véronique FERRAND-RIGALLAUD, C.R.A. (Centre Hospitalier Universitaire de Poitiers, Direction de la Recherche Clinique, Poitiers, France.), Séverine CLERJAUD, (Centre Hospitalier Universitaire de Poitiers, Direction de la Recherche Clinique, Poitiers, France.), Flora DJANIKIAN, M.D. (Centre Hospitalier Universitaire de Montpellier, Anaesthesia and Intensive Care Department, Montpellier, France.), Pierre-François PERRIGAULT, M.D. (Centre Hospitalier Universitaire de Montpellier, Anaesthesia and Intensive Care Department, Montpellier, France.), Karim LAKKHAL, M.D. (Nantes Université, Centre Hospitalier Universitaire de Nantes, Service d’Anesthésie Réanimation, F-44000, Nantes, France.), Matthieu BIAIS, M.D., Ph.D. (Centre Hospitalier Universitaire de Bordeaux, Anaesthesia and Intensive Care Unit, Bordeaux, France), Grégoire CHADEFAUX, M.D. (Centre Hospitalier Universitaire de Bordeaux, Anaesthesia and Intensive Care Unit, Bordeaux, France) Aurore RODRIGUES, M.D. (Centre Hospitalier Universitaire du Kremlin-Bicêtre, Paris, France), Jean-François PAYEN, M.D., PhD (Centre Hospitalier Universitaire de Grenoble, Anaesthesia and Intensive Care Department, Grenoble, France), Magdalena SZCZOT M.D. (Centre Hospitalier Universitaire de Strasbourg, Anaesthesia and Intensive Care Department, Strasbourg, France), Stéphane HECKETSWEILER C.R.A. (Centre Hospitalier Universitaire de Strasbourg, Direction de la Recherche Clinique, Strasbourg, France.), Nicolas ENGRAND M.D.(Hôpital Fondation Adolphe de Rotchild, Anaesthesia and Intensive Care Unit, Paris, France), Michel WOLFF, M.D, PhD. (Hôpital Sainte Anne, Service de Neuroréanimation, Paris, France), Gérard AUDIBERT, M.D, PhD. (Centre Hospitalier de Nancy, Service d’Anesthésie Réanimation, Nancy, France), Frédéric DAILLER, M.D. (Hospices civils de Lyon, Groupement Est, Service d’Anesthésie Réanimation, Lyon, France), Belaid BOUHEMAD, M.D.,PhD (Service d’anesthésie réanimation médecine périopératoire, Chu Dijon – Dijon, France).

## References

1. Luque-Paz D, Revest M, Eugène F. Ventriculitis: A Severe Complication of Central Nervous System Infections. Open Forum Infect Dis 2021;8(6).

2. Srihawan C, Castelblanco RL, Salazar L. Clinical Characteristics and Predictors of Adverse Outcome in Adult and Pediatric Patients With Healthcare-Associated Ventriculitis and Meningitis. Open Forum Infect Dis 2016;3(2).

3. Martin RM, Zimmermann LL, Huynh M, Polage CR. Diagnostic Approach to Health Care-and Device-Associated Central Nervous System Infections. J Clin Microbiol 2018;56(11).

4. Ramanan M, Lipman J, Shorr A, Shankar A. A meta-analysis of ventriculostomy-associated cerebrospinal fluid infections. BMC Infect Dis 2015;15(3).

5. Tunkel AR, Hasbun R, Bhimraj A. Infectious Diseases Society of America’s Clinical Practice Guidelines for Healthcare-Associated Ventriculitis and Meningitis. Clin Infect Publ Infect Soc Am 2017;2017;64(6):e34–65.

6. Jalusic KO, Hempel G, Arnemann P-H. Population pharmacokinetics of vancomycin in patients with external ventricular drain-associated ventriculitis. Br J Clin Pharmacol 2021;87(6):2502–10.

7. The European Committee on Antimicrobial Susceptibility Testing. Breakpoint tables for interpretation of MICs and zone diameters [Internet]. 2024;Available from: http://www.eucast.org

8. Hashemian SMR, Farhadi T, Ganjparvar M. Linezolid: a review of its properties, function, and use in critical care. Drug Devel Ther 2018;12:1759–67.

9. Sun F, Ruan Q, Wang J. Linezolid manifests a rapid and dramatic therapeutic effect for patients with life-threatening tuberculous meningitis. Antimicrob Agents Chemother 2014;58(10):6297–301.

10. Smith AGC, Gujabidze M, Avaliani T. Clinical outcomes among patients with tuberculous meningitis receiving intensified treatment regimens. Int J Tuberc Lung J Int Union Tuberc Lung Dis 2021;25(8):632–9.

11. Kempker RR, Smith AGC, Avaliani T. Cycloserine and Linezolid for Tuberculosis Meningitis: Pharmacokinetic Evidence of Potential Usefulness. Clin Infect Dis 2022;75(4):682–9.

12. Viaggi B, Cangialosi A, Langer M. Tissue Penetration of Antimicrobials in Intensive Care Unit Patients: A Systematic Review-Part II. Antibiot Basel Switz 2022;11(9).

13. Luque S, Grau S, Alvarez-Lerma F. Plasma and cerebrospinal fluid concentrations of linezolid in neurosurgical critically ill patients with proven or suspected central nervous system infections. Int J Antimicrob Agents 2014;44(5):409–15.

14. Beer R, Engelhardt KW, Pfausler B. Pharmacokinetics of Intravenous Linezolid in Cerebrospinal Fluid and Plasma in Neurointensive Care Patients with Staphylococcal Ventriculitis Associated with External Ventricular Drains. Antimicrob Agents Chemother 2007;51(1):379–82.

15. Myrianthefs P, Markantonis SL, Vlachos K. Serum and cerebrospinal fluid concentrations of linezolid in neurosurgical patients. Antimicrob Agents Chemother 2006;50(12):3971–6.

16. Wu X, Tang Y, Zhang X, Wu C, Kong L. Pharmacokinetics and pharmacodynamics of linezolid in plasma/cerebrospinal fluid in patients with cerebral hemorrhage after lateral ventricular drainage by Monte Carlo simulation. Drug Devel Ther 2018;12:1679–84.

17. Zhao W, Kong L, Wu C, Wu X. Prolonged infusion of linezolid is associated with improved pharmacokinetic/pharmacodynamic (PK/PD) profiles in patients with external ventricular drains. Eur J Clin Pharmacol 2020;

18. Viaggi B, Paolo AD, Danesi R. Linezolid in the central nervous system: comparison between cerebrospinal fluid and plasma pharmacokinetics. Scand J Infect Dis 2011;43(9):721–7.

19. Reiber H, Felgenhauer K. Protein transfer at the blood cerebrospinal fluid barrier and the quantitation of the humoral immune response within the central nervous system. Clin Chim Acta 1987;163(3):319–28.

20. Reiber H. Dynamics of brain-derived proteins in cerebrospinal fluid. Clin Chim Acta Int J Clin Chem 2001;310(2):173–86.

21. Rayner CR, Forrest A, Meagher AK, Birmingham MC, Schentag JJ. Clinical Pharmacodynamics of Linezolid in Seriously Ill Patients Treated in a Compassionate Use Programme. Clin Pharmacokinet 2003;42(15):1411–23.

22. The European Committee on Antimicrobial Susceptibility Testing. Antimicrobial wild type distributions of microorganisms [Internet]. 2024;Available from: https://mic.eucast.org/search/

23. Mouton JW, Dudley MN, Cars O, Derendorf H, Drusano GL. Standardization of pharmacokinetic/pharmacodynamic (PK/PD) terminology for anti-infective drugs: an update. J Antimicrob Chemother 2005;55(5):601–7.

24. Kumta N, Roberts JA, Lipman J, Wong WT, Joynt GM, Cotta MO. A Systematic Review of Studies Reporting Antibiotic Pharmacokinetic Data in the Cerebrospinal Fluid of Critically Ill Patients with Uninflamed Meninges. Antimicrob Agents Chemother 2020;65(1):e01998–20.

25. Yogev R, Damle B, Levy G, Nachman S. Pharmacokinetics and Distribution of Linezolid in Cerebrospinal Fluid in Children and Adolescents. Pediatr Infect J 2010;29(9):827–30.

26. Yekani M, Memar MY. Immunologic biomarkers for bacterial meningitis. Clin Chim Acta 2023;548:117470.

27. König C, Grensemann J, Czorlich P, Schlemm E, Kluge S, Wicha SG. A dosing nomograph for cerebrospinal fluid penetration of meropenem applied by continuous infusion in patients with nosocomial ventriculitis. Clin Microbiol Infect Off Publ Eur Soc Clin Microbiol Infect Dis 2022;28(7):1022.e9–1022.e16.

28. Nau R, Lassek C, Kinzig-Schippers M, Thiel A, Prange HW, Sörgel F. Disposition and elimination of meropenem in cerebrospinal fluid of hydrocephalic patients with external ventriculostomy. Antimicrob Agents Chemother 1998;42(8):2012–6.

29. Ishikawa M, Yamazaki S, Suzuki T, Uchida M, Iwadate Y, Ishii I. Correlation between vancomycin penetration into cerebrospinal fluid and protein concentration in cerebrospinal fluid/serum albumin ratio. J Infect Chemother J Jpn Soc Chemother 2019;25(2):124–8.

30. Tarnaris A, Toma AK, Chapman MD. Rostrocaudal dynamics of CSF biomarkers. Neurochem Res 2011;36(3):528–32.

31. Weisner B, Bernhardt W. Protein fractions of lumbar, cisternal, and ventricular cerebrospinal fluid. Sep Areas Ref J Neurol Sci 1978;37(3):205–14.

32. Chen C, Xu P, Xu T, Zhou K, Zhu S. Influence of cerebrospinal fluid drainage and other variables on the plasma vancomycin trough levels in postoperative neurosurgical patients. Br J Neurosurg 2021;35(2):133–8.

33. Li X, Wang X, Wu Y, et al. Plasma and cerebrospinal fluid population pharmacokinetic modeling and simulation of meropenem after intravenous and intrathecal administration in postoperative neurosurgical patients. Diagn Microbiol Infect Dis 2019;93(4):386–92.

34. Andes D, van Ogtrop ML, Peng J, Craig WA. In Vivo Pharmacodynamics of a New Oxazolidinone (Linezolid). Antimicrob Agents Chemother 2002;46(11):3484–9.

35. Fu X, Lin Z, Chen S, Hong L, Yu X, Wu S. Treatment of Intracranial Infection Caused by Methicillin-Resistant Staphylococcus epidermidis with Linezolid Following Poor Outcome of Vancomycin Therapy: A Case Report and Literature Review. Infect Drug Resist 2021;14:2533–42.

36. Shi C, Xia J, Ye J. Effect of renal function on the risk of thrombocytopaenia in patients receiving linezolid therapy: A systematic review and meta-analysis. Br J Clin Pharmacol 2022;88(2):464–75.

37. Crass RL, Cojutti PG, Pai MP, Pea F. Reappraisal of Linezolid Dosing in Renal Impairment To Improve Safety. Antimicrob Agents Chemother 2019;63(8).

38. Cattaneo D, Orlando G, Cozzi V, et al. Linezolid plasma concentrations and occurrence of drug-related haematological toxicity in patients with Gram-positive infections. Int J Antimicrob Agents 2013;41(6):586–9.

39. Wang X, Wang Y, Yao F. Pharmacokinetics of Linezolid Dose Adjustment for Creatinine Clearance in Critically Ill Patients: A Multicenter, Prospective, Open-Label, Observational Study. Drug Devel Ther 2021;15:2129–41.

40. Delattre IK, Taccone FS, Jacobs F, et al. Optimizing β-lactams treatment in critically-ill patients using pharmacokinetics/pharmacodynamics targets: are first conventional doses effective? Expert Rev Anti Infect Ther 2017;15(7):677–88.

