## Supplemental materials for "CSF pharmacokinetics-pharmacodynamics of linezolid in critically brain injured patients, with or without central nervous system healthcare-associated infection. The PK-Pop-LCR Study: A Multicenter Pharmacokinetics and Pharmacodynamics Population Study"

*Supplementary Appendix*

**PK-Pop-LCR Study**

This appendix has been provided by the authors to give readers additional information about their work.

__Table of Contents__


**PK-Pop-LCR STUDY GROUP**

**MATERIAL AND METHODS**

**Linezolid assay**

**Pharmacokinetic analysis**

Structural model

Statistical model

Covariate analysis

Model discrimination and evaluation

Secondary parameters

**Figure S1.** Schematic representation of the final PK model. V1, $V_{CSF}$ and $V_{EVD}$ correspond to volumes of central compartment, CSF compartment and CSF samples collected via the EVD, respectively; $Q_{CSF,in}$ and $Q_{CSF,out}$ are the CSF input and output clearances respectively; $CL_{EVD}$ represents the EVD flow rate fixed for each patient at the experimental value obtained for each sampling time interval; $R_{inf}$ is the rate of infusion of linezolid.

**RESULTS**

**Table S1 -** Patients demographics and characteristics

**Table S2**. Parameters of the pharmacokinetic model

**Table S3.** Cumulative fraction of response (CFR) of linezolid against methicillin resistant *S. aureus* (MRSA) and *S. epidermidis* (MRSE) in simulated patients receiving different daily doses considering a CSF PK/PD target of 100% T>MIC.

**Figure S2.** Boxplot of the CSF/plasma albumin ratio ($Q_{alb}$) in patients with "Extra-CNS Infection" (blue) and those with "CNS Infection" (yellow).

**Figure S3.** Visual predictive check of the pharmacokinetic model for total plasma and unbound CSF concentrations. Circles represent observed concentrations, solid lines the medians of the 1000 simulations and the blue area depict the 90% prediction interval for those simulations.

**Figure S4.** Individual predictions (solid line) and measured concentrations (points) of linezolid in plasma (red) and $CSF_{EVD}$ (blue).

**Figure S5.** Goodness of fit (GOF) plots for the pharmacokinetic model. A: observed data versus population predicted concentrations in plasma (red) and in $CSF_{EVD}$ (blue). B: observed data versus individual predicted concentrations in plasma (red) and in $CSF_{EVD}$ (blue). C: conditional weighted residuals (CWRES) versus population predictions in plasma (red) and in $CSF_{EVD}$ (blue). D: Conditional weighted residuals versus time for plasma (red) and $CSF_{EVD}$ (blue).

**Figure S6:** Probability of achieving the PK/PD target of 100% *f*T > MIC in CSF (solid lines) in simulated patients receiving the same daily dose of linezolid (1 800 mg/day) but divided into 2 doses (red), 3 doses (green) or administered as a continuous infusion (blue). The distribution of linezolid MICs for methicillin resistant *S. aureus* (MRSA) and *S. epidermidis* (MRSE) is shown.

**REFERENCES**

**PK-Pop-LCR STUDY GROUP**

**List of collaborators for each center**

**CHU La Miletrie, Poitiers, France:** Fanny BERNARD, M.D., Carine DOMINIQUE, M.D., Amélie RUBIN, M.D., Tony TRIQUARD, M.D., Aude PERALEZ, M.D., Adriana DELWAIL, L.E., Rémy BELLIER, M.D., Thierry BENARD, M.D., Nadia IMZI, C.R.A., Angela KOSTENCOVSKA, C.R.A., Sabrina SEGUIN, C.R.A., Véronique FERRAND-RIGALLAUD, C.R.P.M, Séverine CLERJAUD, D.M.

**CHU de Montpellier, Montpellier, France :** Flora Djanikian, M.D., Pierre-François Perrigault, P.D.,

**CHU de Bordeaux, Bordeaux, France :** Matthieu Biais, M.D., PhD, Grégoire Chadefaux, M.D.

**CHU du Kremlin-Bicêtre, Paris, France :** Aurore Rodrigues, M.D.

**CHU de Grenoble, Grenoble, France :** Jean-François Payen, M.D., PhD.

**CHU de Strasbourg, Strasbourg, France :** Magdalena Szczot M.D., Stéphane Hecketsweiler C.R.A.

**Hôpital Fondation Adolphe de Rotchild, Paris, France :** Nicolas Engrand M.D.

### MATERIAL AND METHODS

#### Linezolid assay

Linezolid standard (purity > 99% in powder form) was purchased from Sigma-Aldrich (Saint-Quentin-Fallavier, France). Linezolid D8 (purity of 100% in powder form) was obtained from Alsachim (Illkirch Graffenstaden, France).

Analysis of linezolid in human plasma and CSF were performed by an LC-MS/MS method, previously described and developed for the quantification of ten antibiotics in human plasma for routine therapeutic drug monitoring[2]. Calibration curves were established in both human plasma and NaCl solution (used for CSF matrix) over 10 to 10000 ng/mL. Plasma samples were precipitated with 100 µL of acetonitrile containing the internal standard (Linezolid-D8) at 100 ng/mL. The tubes were then vortexed for 30 s and centrifuged at 17,160×g for 5 min at 4 °C. A volume of 100 µL of supernatant was transferred into a glass tube to which 100 µL of 10 mM ammonium formate were added prior to injection. For the CSF samples, a 1/200 dilution in a methanol/water mixture (50/50, v/v) containing the internal standard at 500 ng/mL was performed. Tubes were then vortexed for 30 s before injection.

The system included a Shimadzu high-performance liquid chromatography system module (Nexera XR; Shimadzu, Marne la Vallée, France) coupled with a TQ3500 mass spectrometer (Sciex, Les Ulis, France). The compound was analyzed on an XBridge Peptide BEH300 C18 column (5µm, 2.1 x 150 mm, Waters, Saint-Qunetin-en Yvelines, France). The mobile phase consisted of a mixture (50/50, v/v) of 10 mM ammonium formate and acetonitrile delivered isocratically at 0.20 mL/min. Electrospray ionization in positive mode was used for the detection. Ions were analyzed in the multiple reaction monitoring, and the following transitions were inspected: m/z 338→296 and 346→304 for linezolid and internal standard, respectively. The intraday variability was characterized at three concentration levels (20, 1000 and 7500 ng/ml for both matrices) with a precision < 8% and bias < 13% for both matrices. Corresponding between-day variability was determined with a precision < 3% and a bias < 7% for both matrices (n = 16 for plasma and n=14 for NaCl matrices).

#### Pharmacokinetic analysis

Structural model

Total plasma linezolid concentrations were first analyzed using a one-compartment model with first-order elimination, parametrized in terms of clearances (CL) and volume of distribution (V1). Plasma parameters estimated and corrected for protein binding of 31%[4] were then fixed to fit CSF data. In the final model (Figure S1), parameters for both plasma and CSF data were estimated simultaneously. The CSF was represented as a compartment characterized by a volume ($V_{CSF}$) which was fixed to a value of 0.15 L corresponding to the physiological volume of cranial CSF[5] due to identifiability issue. The bidirectional passage across the BCSFB was characterized by a clearance into the CSF ($Q_{CSF,in}$) and a clearance out of the CSF ($Q_{CSF,out}$). Since CSF data were obtained from the EVD, a third compartment was added into the model. Linezolid concentrations in the EVD compartment were modelled for each collection interval using the flow rate of the EVD ($CL_{EVD}$) and the volume of CSF samples ($V_{EVD}$) determined experimentally as covariates. When actual sampling times were missing, theoretical times were associated to the samples.

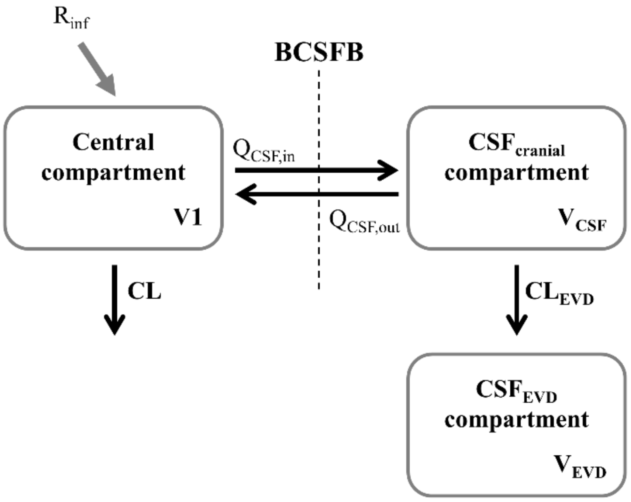


**Figure S1.** Schematic representation of the final PK model. V1, $V_{CSF}$ and $V_{EVD}$ correspond to volumes of central compartment, CSF compartment and CSF samples collected via the EVD, respectively; $Q_{CSF,in}$ and $Q_{CSF,out}$ are the CSF input and output clearances respectively; $CL_{EVD}$ represents the EVD flow rate fixed for each patient at the experimental value obtained for each sampling time interval; $R_{inf}$ is the rate of infusion of linezolid.

Statistical model

Inter-individual variations in PK parameters was best described by an exponential model and the residual variability, resulting from unexplained sources, was best described with proportional error models for total plasma and unbound CSF linezolid concentrations.

Covariate analysis

The influence of various patient characteristics on the estimates of PK parameters were investigated with the stepwise covariate modeling (SCM) procedure implemented in PsN (version 4.8.1), using a forward inclusion (p-value <0.01) followed by backward elimination process (p-value <0.001). Linear, power and exponential functions were evaluated for all tested covariates. Covariates tested included weight, age, height, creatinine clearance, serum and CSF albumin, total protein, glucose and inflammatory biomarkers (IL6, IL8, IL10, IL17A, TNF-alpha and IgG). Covariates were normalized by the corresponding median value of the individuals studied. When no value of a continuous covariate was available for one individual, the median value calculated for the PK population was used, and for a categorical covariate, it was replaced by the mode of this covariate. The evaluation of the covariate model was based on relevance of the covariate on the parameter, improvement of diagnostic plots, parameter uncertainty of the additional parameter and, if applicable, reduction of inter-individual variability in the parameter concerned.

Model discrimination and evaluation

Discrimination between models and model evaluation were based on the visual inspection of diagnostic plots - goodness of fit (GOF) and individual plots - comparison of the objective function value (OFV) and relative standard errors (RSEs) of the parameter estimates. RSEs of parameters were obtained using the Sampling Importance Resampling (SIR) procedure[6] implemented in PsN (version 4.8.1).

A simulation-based method (visual predictive checks – VPCs) was used to evaluate the adequacy of the final model. The final PK model estimates were used to simulate 1000 concentrations-time profiles with the same study design characteristics as the original dataset. The $5^{th}$, $50^{th}$ (median) and the $95^{th}$ percentiles of the simulated concentrations and raw data were calculated and plotted versus time since last dose, the concordance between simulations and observations was inspected visually.

Secondary parameters

Following secondary parameters were estimated from Monte-Carlo simulations: the free maximal concentrations ($C_{max}$) and the free area under the curve of concentrations from 0 to 24 h at steady-state ($fAUC_{0-24}$) for plasma and CSF. The secondary parameters were estimated from 5 000 concentration-time profiles simulated with the final model after administration of 600 mg q12h and 600 mg q8h, using as input a dataset containing 25 individuals with the same covariate values as the original dataset.

**RESULTS**

**Table S1 -** Patients demographics and characteristics

| Patient Identity | Gender | Body Weight (kg) | Height (cm) | $CL_{cr}$ (mL/min) (MDRD) | $CL_{cr}$ (mL/min) (measured) | Mean $CL_{EVD}$ (mL/h) | SOFA score | SAPS II score | Leucocytes in plasma ($10^9$/L) | Lactates in CSF (mmoL/L) | Protein in CSF (g/L) | CSF/blood glucose ratio | Infection diagnosed * | Infectious agents in CNS | Linezolid MIC (mg/L) |
|---|---|---|---|---|---|---|---|---|---|---|---|---|---|---|---|
| 1 | F | 50 | 162 | 206 | 195 | 12.3 | 6 | 24 | 16.2 | 2.00 | 0.17 | 0.34 | Systemic | -- | 2 |
| 2 | F | 96 | 163 | 180 | 216 | 12.6 | 3 | 39 | 14.3 | n.a. | n.a. | n.a. | Systemic | -- | 2 |
| 3 | M | 81 | 170 | 76 | 119 | 20.2 | 0 | 50 | 13.3 | n.a. | n.a. | n.a. | CNS | *S. epidermidis* | 1 |
| 4 | M | 140 | 185 | 118 | 171 | 14.1 | 10 | 56 | 11.8 | 3.40 | 2.23 | 0.61 | Systemic | -- | n.a. |
| 5 | M | 71 | 165 | 259 | 87 | 9.06 | 5 | 51 | 12.4 | 5.00 | 1.20 | 0.41 | CNS | *S. epidermidis* | 1 |
| 6 | F | 72 | 165 | 123 | 128 | 2.64 | 2 | 37 | 5.60 | 2.40 | 1.00 | 0.54 | CNS | *S. epidermidis* | 2 |
| 7 | F | 59 | 166 | 418 | 166 | 9.38 | 8 | 29 | 4.40 | 10.3 | 0.63 | 0.18 | CNS | *S. aureus* | 2 |
| 8 | M | 120 | 171 | 381 | 246 | 10.3 | 5 | 34 | 7.50 | 2.30 | 0.41 | 0.47 | Systemic | -- | 2 |
| 9 | F | 47 | 153 | 228 | 105 | 15.1 | 2 | 29 | 9.80 | 2.80 | 0.30 | 0.37 | CNS | *S. pettenkoferi* | 1 |
| 10 | F | 60 | 157 | 380 | 335 | 12.1 | 8 | 49 | 8.10 | 6.40 | 0.37 | 0.50 | CNS | *S. epidermidis* | 1 |
| 11 | M | 88 | 175 | 182 | 147 | 10.2 | 3 | 30 | 24.3 | 3.80 | 0.60 | 0.40 | Systemic | -- | 2 |
| 12 | M | 115 | 188 | 120 | 176 | 6.30 | 2 | 50 | 11.8 | 1.90 | 0.24 | n.a. | Systemic | -- | 2 |
| 13 | M | 93 | 165 | 106 | 65 | 11.9 | 5 | 31 | 9.24 | n.a. | n.a. | n.a. | Systemic | -- | n.a. |
| 14 | F | 75 | 162 | n.a. | n.a. | 12.2 | 1 | 45 | n.a. | 5.30 | 0.34 | n.a. | Systemic | -- | n.a. |
| 15 | F | 80 | 163 | 273 | 88 | 7.04 | 3 | 32 | 9.90 | 5.90 | 0.28 | 0.31 | CNS | *M. tuberculosis* | n.a. |
| 16 | M | 90 | 175 | 188 | n.a. | 2.51 | 1 | 29 | 8.60 | n.a. | 0.50 | n.a. | CNS | Flora of Gram-negative bacillus | n.a. |
| 17 | M | 74 | 175 | 183 | 49 | 4.39 | 0 | 20 | 8.60 | n.a. | 0.21 | 0.64 | CNS | *S. epidermidis* | 0.75 |
| 18 | F | 43 | 161 | 244 | 184 | 15.6 | 2 | 43 | 14.6 | 2.30 | 0.57 | 0.59 | CNS | *C. acnes* | 0.25 |
| 19 | F | 70 | 173 | 115 | n.a. | 9.00 | 6 | 59 | 23.1 | 3.40 | 0.76 | 0.58 | Systemic | -- | n.a. |
| 20 | F | 58 | 175 | 273 | n.a. | 19.4 | 3 | 32 | 10.4 | 2.40 | 0.13 | n.a. | CNS | *C. acnes* | 0.5 |
| 21 | M | 88 | 175 | 591 | 281 | 5.27 | 5 | 28 | 7.80 | 4.00 | 0.36 | 0.48 | CNS | *S. epidermidis* | 1.5 |
| 22 | F | 67 | 165 | 129 | n.a. | 16.8 | 1 | 33 | 7.90 | n.a |  | n.a. | CNS | *C. acnes* | n.a. |
| 23 | M | 60 | 157 | n.a. | n.a. | 5.56 | 1 | 19 | 7.40 | 3.20 | 0.60 | n.a. | CNS | *S. epidermidis* | n.a. |
| 24 | F | 51 | 169 | 154 | n.a. | 5.44 | 1 | 31 | 15.8 | n.a. | n.a. | n.a. | Systemic | -- | n.a. |
| 25 | F | 91 | 150 | 70 | n.a. | 5.81 | 8 | 52 | 13.0 | n.a. | n.a. | n.a. | Systemic | -- | n.a. |
| **Median** |  | 74 | 165 | 183 | 166 | 10.2 | 3 | 33 | 10.2 | 3.40 | 0.41 | 0.48 |  |  |  |
| **Quartiles** |  | 60-90 | 162-175 | 122-266 | 105-195 | 5.81-12.6 | 1-5 | 29-49 | 8.05-13.6 | 2.40-5.00 | 0.29-0.62 | 0.38-0.57 |  |  |  |
| **n** | 11 M<br>14 F |  |  |  |  |  |  |  |  |  |  |  | 14 CNS *<br>11 Systemic |  |  |
| **%** | 44% M<br>56% F |  |  |  |  |  |  |  |  |  |  |  | 56% CNS<br>44% Systemic |  |  |

M: male, F: female, $CL_{cr}$: creatinine clearance, $CL_{EVD}$: flow rate of the external ventricular drain, SOFA: Sequential Organ Failure Assessment, SAPS II: Simplified Acute Physiology Score II
n.a: Not Available. * After adjudication : 9 associated-healthcare meningitis and 5 ventriculitis

Table S2. Parameters of the pharmacokinetic model

| Parameter | Definition | Estimate | 95% CI |
|---|---|---|---|
| $V1_{pop}$ (L) | Typical volume of the central systemic compartment | 36.4 | [32.5 – 41.9] |
| $\theta_{V1}$ | Coefficient of the linear relationship between $V1_{pop}$ and patient weight | 0.0115 | [ 0.00622 – 0.0168] |
| $V1=V1_{pop}\times\left(1+\theta_{V1}\times(WT\text{-}74)\right)$ | Relationship between $V1_{pop}$ and patient weight (WT) | | |
| CL (L/h) | Plasma clearance | 8.38 | [7.00 – 9.93] |
| $V_{CSF}$ (L) | Volume of CSF compartment | 0.15 fixed | |
| $Q_{CSF,in}$ (L/h) | CSF input clearance | 0.0400 | [0.0330 – 0.0478] |
| $Q_{CSF,out}$ (L/h) | CSF output clearance | 0.0367 | [0.0298 – 0.0437] |
| **Inter-individual variability (IIV)** | | | |
| $\omega^2_{V1}$ (CV%) | IIV in V1 | 23.1 | [13.9 – 34.2] |
| $\omega^2_{CL}$ (CV%) | IIV in Cl | 45.4 | [35.2 – 59.9] |
| $\omega^2_{QCSF,in}$ (CV%) | IIV in $Q_{CSF,in}$ | 15.0 | [6.24 – 23.8] |
| **Residual Variability (RV)** | | | |
| $\sigma_{plasma}$ (%) | Proportional RV for plasma | 22.1 | [18.2 – 27.1] |
| $\sigma_{CSF}$ (%) | Proportional RV for CSF | 32.6 | [28.6 – 37.5] |

Table S3. Cumulative fraction of response (CFR) of linezolid against methicillin resistant *S. aureus* (MRSA) and *S. epidermidis* (MRSE) in simulated patients receiving different daily doses considering a CSF PK/PD target of 100% T>MIC.

| | 600mg q12h | 600mg q8h | 600mg q6h | 900mg q12h | 1200mg/day continuous infusion | 1800mg/day continuous infusion | 2400mg/day continuous infusion |
|---|---|---|---|---|---|---|---|
| MRSA | 32% | 70% | 88% | 50% | 70% | 88% | 95% |
| MRSE | 74% | 97% | 99% | 86% | 99% | 100% | 100% |

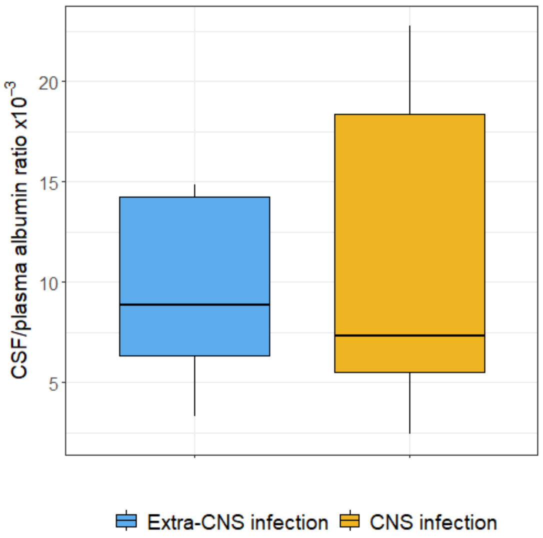


**Figure S2.** Boxplot of the CSF/plasma albumin ratio ($Q_{alb}$) in patients with “Extra-CNS Infection” (blue) and those with “CNS Infection” (yellow).

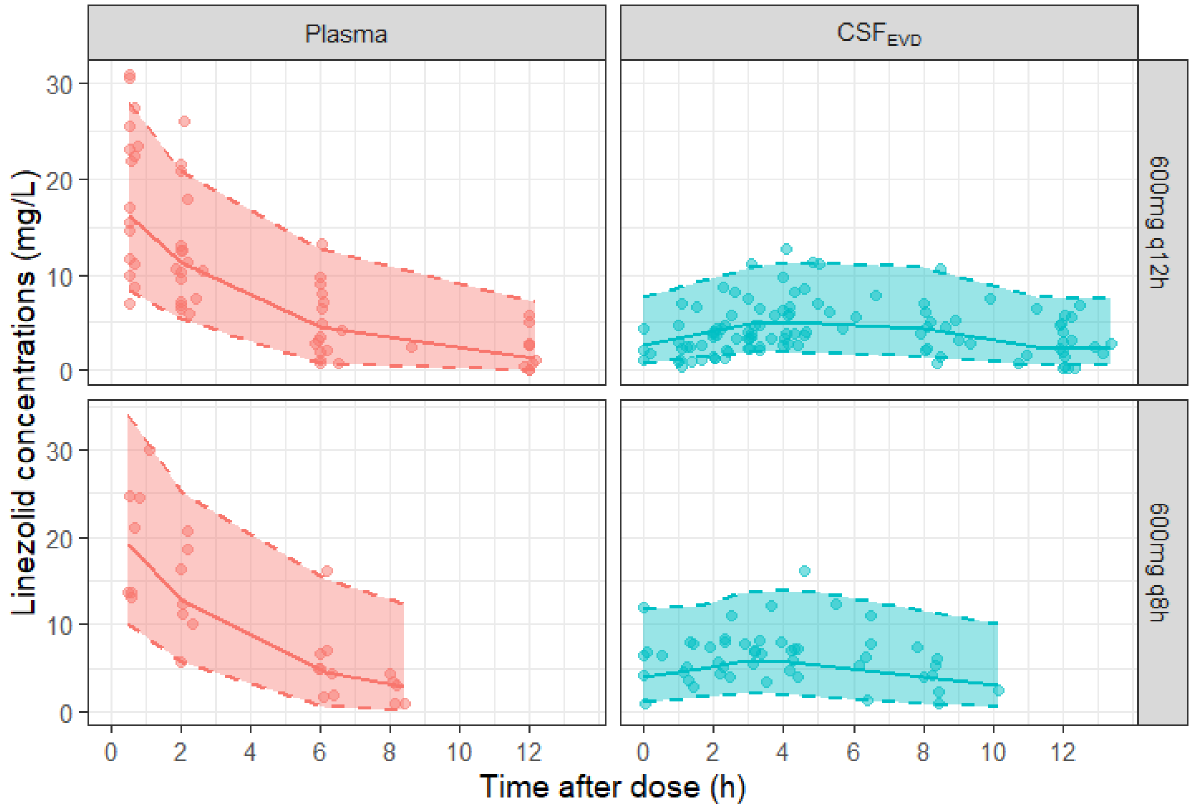


**Figure S3.** Visual predictive check of the pharmacokinetic model for total plasma and unbound CSF concentrations. Circles represent observed concentrations, solid lines the medians of the 1000 simulations and the blue area depict the 90% prediction interval for those simulations.

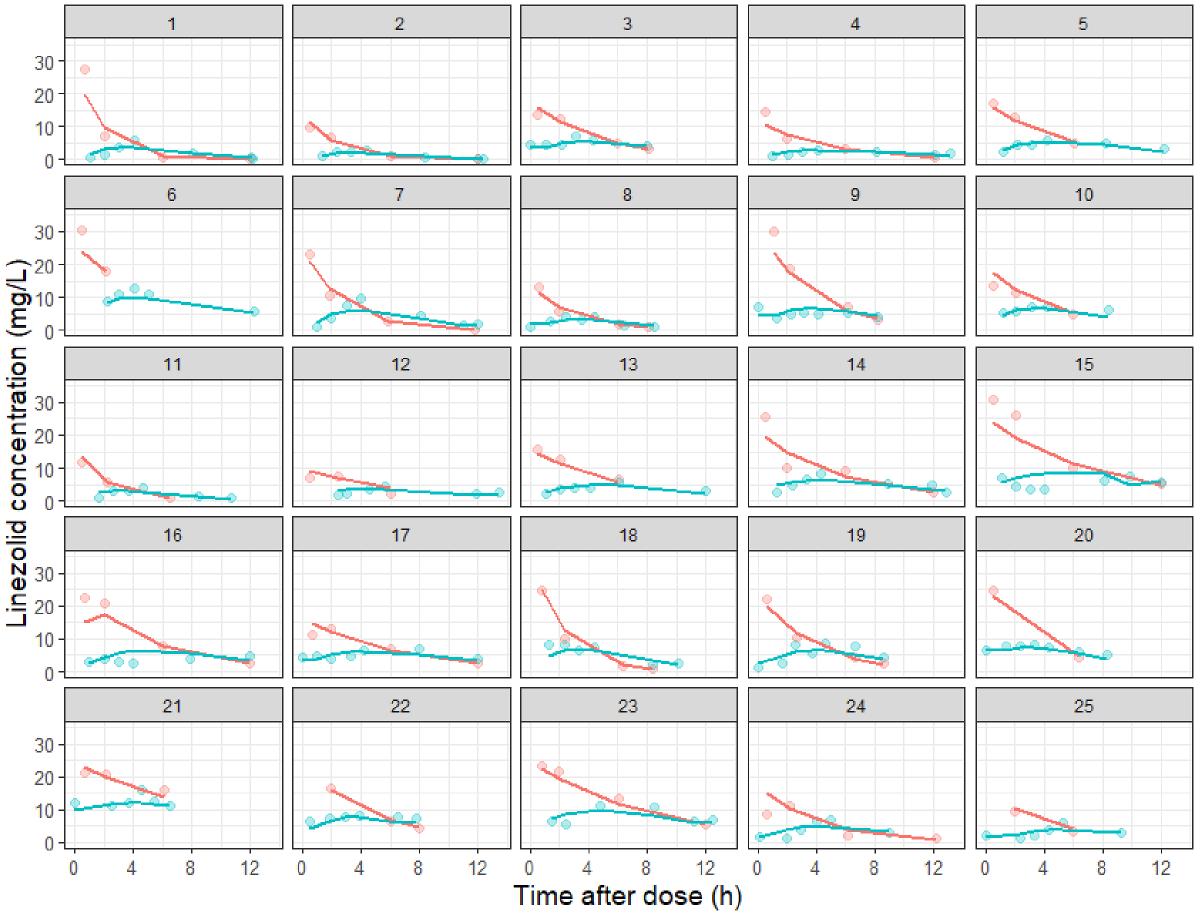


**Figure S4.** Individual predictions (solid line) and measured concentrations (points) of linezolid in plasma (red) and $CSF_{EVD}$ (blue).

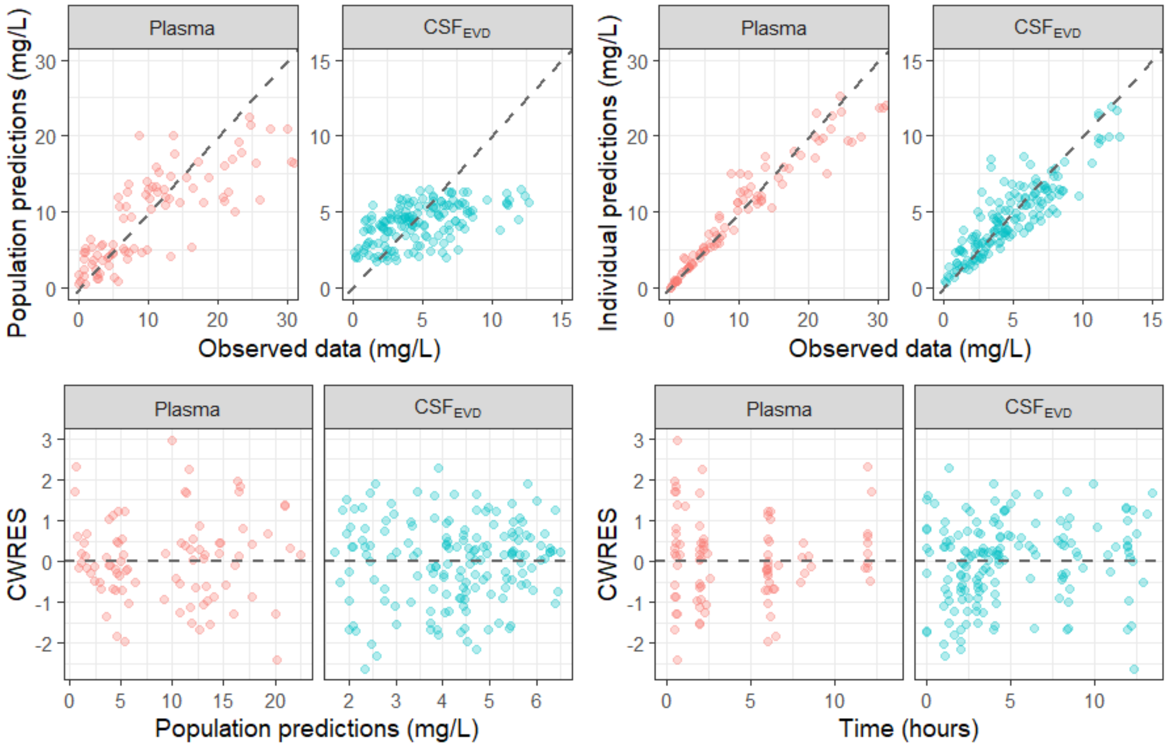


**Figure S5.** Goodness of fit (GOF) plots for the pharmacokinetic model. A: observed data versus population predicted concentrations in plasma (red) and in CSF$_{EVD}$ (blue). B: observed data versus individual predicted concentrations in plasma (red) and in CSF$_{EVD}$ (blue). C: conditional weighted residuals (CWRES) versus population predictions in plasma (red) and in CSF$_{EVD}$ (blue). D: Conditional weighted residuals versus time for plasma (red) and CSF$_{EVD}$ (blue).

**Figure S6.** Probability of achieving the PK/PD target of 100% *f*T > MIC in CSF (solid lines) in simulated patients receiving the same daily dose of linezolid (1 800 mg/day) but divided into 2 doses (red), 3 doses (green) or administered as a continuous infusion (blue). The distribution of linezolid MICs for methicillin resistant *S. aureus* (MRSA) and *S. epidermidis* (MRSE) is shown.

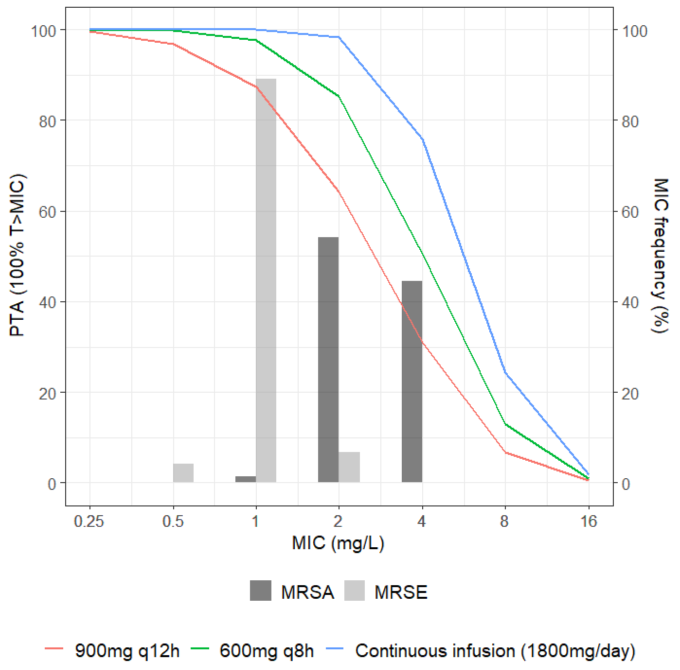

**REFERENCES**


1. Tunkel AR, Hasbun R, Bhimraj A. Infectious Diseases Society of America's Clinical Practice Guidelines for Healthcare-Associated Ventriculitis and Meningitis. Clin Infect Dis Off Publ Infect Dis Soc Am 2017;2017;64(6):e34–65.

2. Radovanovic M, Day RO, Jones GDR, Galettis P, Norris RLG. LC-MS/MS method for simultaneous quantification of ten antibiotics in human plasma for routine therapeutic drug monitoring. J Mass Spectrom Adv Clin Lab 2022;26:48–59.

3. Reiber H, Felgenhauer K. Protein transfer at the blood cerebrospinal fluid barrier and the quantitation of the humoral immune response within the central nervous system. Clin Chim Acta 1987;163(3):319–28.

4. Hashemian SMR, Farhadi T, Ganjparvar M. Linezolid: a review of its properties, function, and use in critical care. Drug Des Devel Ther 2018;12:1759–67.

5. de Lange ECM. Utility of CSF in translational neuroscience. J Pharmacokinet Pharmacodyn 2013;40(3):315–26.

6. Dosne A-G, Bergstrand M, Harling K, Karlsson MO. Improving the estimation of parameter uncertainty distributions in nonlinear mixed effects models using sampling importance resampling. J Pharmacokinet Pharmacodyn 2016;43(6):583–96.
